# Disease-Associated HLA Alleles and Haplotypes in Multiple Sclerosis: Insights from an Albanian Case–Control Study

**DOI:** 10.64898/2025.12.01.25341397

**Authors:** Genc Sulcebe, Erkena Shyti, Margarita Kurti, Donika Kasemi, Armanda Deda, Ilia Mikerezi, Rozarta Nezaj, Ariola Verlaku, Sara Likaj, Jera Kruja

## Abstract

**Background:** Multiple sclerosis (MS) is a complex autoimmune disease with the HLA region showing the strongest known genetic influence. Although HLA-DRB1*15:01 is a well-known risk allele in European populations, data from Southeastern Europe—and Albania specifically—are still limited.

**Objective:** To assess allele- and haplotype-level associations at HLA class I (A*, B*, C*) and class II (DRB1*, DQB1*) loci with MS risk in Albanians, and to explore their correlations with clinical phenotypes.

**Methods:** We conducted a case–control study involving 128 consecutive Albanian MS patients diagnosed according to the 2017 revised McDonald criteria and 148 unrelated healthy Albanian controls. Second-field HLA genotyping (PCR-SSP) was performed at A*, B*, C*, DRB1*, and DQB1* loci. Allele frequencies were calculated through direct gene counting, and Hardy–Weinberg equilibrium (HWE) was tested for each locus. Frequencies of five-locus haplotypes (A∼B∼C∼DRB1∼DQB1) were estimated using maximum likelihood with an expectation–maximization (EM) algorithm. Differences between groups were evaluated with Fisher’s exact test and odds ratios (OR), along with 95% confidence intervals (CI). Multiple testing corrections were applied using the Bonferroni and Benjamini–Hochberg false discovery rate (FDR) methods. Clinical correlations were compared between DRB1*15:01 carriers and non-carriers across phenotype categories (RR, SP, PP), EDSS scores, relapse rates, and MRI activity.

**Results:** All loci in controls conformed to HWE; in patients, class I loci conformed while DRB1* and DQB1* showed deviations. Carrier status for HLA-DRB1*15:01 was significantly higher in patients (40.6%) compared to controls (16.2%) (OR = 3.53; p < 0.0001), remaining significant after Bonferroni and FDR corrections. HLA-DQB1*06:02 also stayed significant after correction, consistent with linkage disequilibrium with DRB115:01. Other significant statistical differences did not persist after multiple testing adjustments. Patients exhibited greater five-locus haplotype diversity than controls (17 vs. 10 haplotypes ≥1%). A patient-enriched haplotype, A*68:01∼B*18:01∼C*07:01∼DRB1*15:01∼DQB1*06:02, was common among cases and absent in controls, whereas A*02:01∼B*51:01∼C*15:02∼DRB1*16:01∼DQB1*05:02 was less frequent in cases and remained protective after Bonferroni correction. No significant differences were seen in clinical course (RR/SP/PP), EDSS measures, relapse rate, or MRI activity between DRB115:01 carriers and non-carriers. At the population level, MS prevalence across 22 European countries was positively correlated with DRB115:01 frequency (Spearman ρ = 0.645; p = 0.0012).

**Conclusions:** In Albanians, HLA-DRB1*15:01 significantly increases the risk of MS, with DQB1*06:02 reflecting the same signal due to strong linkage. Risk- and protection-associated five-locus haplotypes further clarify the HLA genetic architecture in MS patients; however, DRB1*15:01 carriage does not noticeably influence the clinical phenotype or the severity of disease outcome. The relatively lower frequency of the DRB1*15:01 allele in the Albanian population may help explain the country’s lower MS prevalence compared to Northern Europe. Larger cohorts and inclusion of non-HLA loci are necessary to identify additional factors contributing to MS susceptibility and progression in this population.

## Introduction

Multiple sclerosis (MS) is the most common chronic inflammatory disease of the central nervous system (CNS), mainly affecting young adults aged 20 to 40 years, with a notable female predominance (female-to-male ratio approximately 2:1) [1, 2]. The cause of MS is multifactorial, involving both genetic and environmental factors that contribute to disease susceptibility [3]. Genetic factors are believed to account for about 50% of the overall risk, while environmental exposures mainly influence the remaining risk [4]. Familial MS accounts for around 12. 6% of all cases, and the risk of developing the disease increases with the degree of genetic relatedness to the affected individual [5].

Geographically, MS has a higher prevalence in Northern Europe, but over the past few decades, the incidence has steadily increased across Southern European countries, likely due to changes in lifestyle and environmental conditions [6, 7, 8]. Several environmental risk factors have been consistently linked to MS, including Epstein–Barr virus infection, low sunlight exposure and vitamin D deficiency, smoking, and obesity [9, 10, 11].

Genetic research has greatly improved our understanding of how MS develops. MS is considered a multifactorial disease, and genome-wide association studies (GWAS) have identified 233 common genetic variants associated with MS [3, 12]. Of these, 32 have been located in the human leukocyte antigen (HLA) region, while 201 are outside this locus [12, 13]. Besides the HLA region, variants in immune-related genes, such as interleukin receptors IL-2 (IL2RA) and IL-7 (IL7R), have also been linked to disease risk [14, 15]. However, the strongest genetic association is found within the major histocompatibility complex (MHC), especially with the HLA-DRB1*15:01* allele, which increases susceptibility approximately threefold in carriers [16, 17].

Given the significant genetic contribution and extensive allelic diversity of the HLA system across populations, exploring HLA-allele associations within specific populations remains highly important. Data on MS genetics in Southeastern Europe, particularly in Albania, are limited. Understanding how HLA alleles are distributed among Albanian MS patients and their potential link to clinical subtypes could offer new insights into disease susceptibility patterns and improve our overall understanding of MS development.

## Materials and Methods

### Study design and populations

#### Multiple sclerosis patients

This study included 128 consecutive individuals with MS at the Neurology Service of the University Hospital Center “Mother Teresa” between 22 July 2024 and 29 May 2025. Written informed consent was obtained from all participants. The diagnosis of MS was established according to the 2017 Revised McDonald clinical and imaging criteria [18].

#### Control group

Control individuals were analyzed to establish the baseline distribution of HLA alleles in the Albanian population. A total of 148 samples were collected from healthy volunteers, including medical students and blood donors at the National Blood Donation Center in Tirana. Eligibility criteria for controls included: (i) no self-reported history of chronic disease, (ii) confirmed Albanian ethnicity for at least three generations, (iii) no kinship with other participants, and (iv) origin from a specific geographical region of Albania.

#### Ethical approval

The study protocol received approval from the Ethics Committee of the Albanian Academy of Sciences (Project No. 373, dated July 15, 2024). All participants provided written informed consent before enrollment.

#### HLA genotyping

Genomic DNA was extracted from the buffy coat of blood samples using the QIAamp DNA Blood Mini Kit (Qiagen, Hilden, Germany). Second-field (allele) level HLA genotyping of class I (HLA-A*, -B*, -C*) and class II (HLA-DRB1*, -DQB1*) loci was performed using PCR with sequence-specific primers (PCR-SSP) and Micro SSP™ DNA Typing Trays (One Lambda, Thermo Fisher Scientific, USA).

#### Data analysis

Allele frequencies were estimated through direct gene counting [19]. Hardy–Weinberg equilibrium (HWE) was tested for each locus (HLA-A*, -B*, -C*, -DRB1*, -DQB1*) using the Arlequin software package [20].

Case–control differences in allele distributions were evaluated using Fisher’s exact test. Odds ratios (OR) with 95% CI were calculated to assess the strength and direction of associations. To account for multiple comparisons, p-values were adjusted with both the Bonferroni correction and the Benjamini–Hochberg false discovery rate (FDR) method [21, 22]. Statistical significance was determined based on the corrected p-values.

Haplotype frequencies were inferred using maximum likelihood estimation with an expectation– maximization (EM) algorithm implemented in the Arlequin program [20]. Comparative data on HLA allele frequencies and MS prevalence in other populations were obtained from published sources [23, 24]. The correlation between allele frequencies and MS prevalence across populations was analyzed using Spearman’s rank correlation test. Differences in categorical and quantitative data between MS patient subgroups were assessed using Fisher’s exact test for categorical data and the Mann-Whitney U test for quantitative data, respectively. All statistical analyses were performed using R software (R Foundation for Statistical Computing, version 4.3.1, R Core Team, 2023, Vienna, Austria).

## Results

### Characteristics of the study sample populations

This study involved 128 consecutive individuals with multiple sclerosis (MS) at the Neurology Service of the University Hospital Center “Mother Teresa” between April 2024 and May 2025. The main demographic and clinical characteristics of the patients are shown in Table 1. Females were more common than males (female-to-male ratio 1.8:1), while age distribution was normal across the entire patient group (Shapiro–Wilk test, p = 0.072). No significant difference was found between the mean age of male patients (35.6 years) and female patients (38.6 years). The control group included 148 unrelated healthy individuals with a mean age of 37.8 years (range, 20-67 years). Among them, 47.3% were female and 52.7% male.

**Table 1.** Demographic and clinical characteristics of patients with multiple sclerosis.

| Demographic and Clinical Data | Values |
| --- | --- |
| Age in years (mean ± SD; 95% CI; range) | 38.0 ± 11.8; 36.0 – 40.1; 15 – 67 |
| Sex (number; %) | *Females:* 82 (64.1 %); *Males:* 46 (35.9 %) |
| Disease duration in years (mean ± SD; 95% CI; range) | 8.5 ± 6.2; 7.4 – 9.5; 0.2 – *28 |
| Residence at disease onset (urban vs rural: number; %) | *Urban:* 103 (80.5 %); *Rural:* 25 (19.5 %) |
| Family history of MS (yes vs no; number; %) | *Yes:* 15 (11.7 %); *No:* 113 (88.3 %) |
| Relapsing - Remitting (RR) clinical form (number; %) | 103 (80.5 %) |
| Secondary - Progressive (SP) clinical form (number; %) | 14 (10.9 %) |
| Primary - Progressive (PP) clinical form (number; %) | 11 (8.6 %) |
| EDSS at diagnosis (points: mean ± SD; 95% CI; range) | 2.07 ± 1.12; 1.87 – 2.27; 0.5 – 6.5 |
| Current EDSS (points: mean ± SD; 95% CI; range) | 2.64 ± 1.88; 2.32 – 2.97; 0 – 7.5 |
| Relapses (during the last 2 years) | 0.64 ± 0.78; 0.50 – 0.78; 0 – 3 |
| MRI with active lesions (number; %) | 45 (35.2 %) |

*EDSS:* Expanded Disability Status Scale.
*RR:* Relapsing-remitting; *SP:* Secondary progressive; *PP:* Primary progressive.
*MRI:* Magnetic Resonance Imaging.

### HLA genotyping results

All 128 MS patients and 148 control individuals were genotyped at the allele level for the HLA-A*, HLA-B*, HLA-C*, HLA-DRB1*, and HLA-DQB1* loci. In the patient group, no deviations from Hardy–Weinberg equilibrium (HWE) were observed for class I loci (HLA-A*, -B*, and -C*), while deviations were noted for DRB1* and DQB1* class II loci. All loci in the control group conformed to HWE.

All allele frequency data and detailed statistical results for both groups are provided in the Supplementary Table (Suppl. Table 1). The comparisons of allele frequencies between patients and controls for alleles with nominal p-values < 0.05 are presented in Table 2. As shown in this table, after applying the Bonferroni and Benjamini-Hochberg corrections for multiple testing, only HLA-DRB1*15:01 and HLA-DQB1*06:02 remained statistically significant. The odds ratios for the significant predisposing and protective alleles are shown in Figure 1.

**Table 2.** Allele frequencies and statistical results for alleles with significant differences between the patient and control groups.

| Predisposing Alleles |  |  |  |  |  | Protective Alleles |  |  |  |  |  |
| --- | --- | --- | --- | --- | --- | --- | --- | --- | --- | --- | --- |
| Allele | Patients<br>(allele<br>frequency<br>in %) | Controls<br>(allele<br>frequency<br>in %) | Fisher<br>test *p*-<br>value<br>(nominal) | Fisher<br>test *p*-<br>value<br>(after<br>Bonferro<br>ni<br>correctio<br>n) | Fisher<br>test *p*-<br>value<br>(after<br>BH-FDR*) | Allele | Patients<br>(allele<br>frequency<br>in %) | Controls<br>(allele<br>frequency<br>in %) | Fisher<br>test *p*-<br>value<br>(nominal) | Fisher test<br>*p*-value<br>(Bonferro<br>ni<br>correction) | Fisher<br>test *p*-<br>value<br>(after<br>BH-FDR*) |
| A*01:01 | 15.55 | 9.68 | 0.0456 | 0.8199 | 0.6250 | B*52:01 | 0.41 | 3.23 | 0.0234 | 0.8184 | 0.6818 |
| B*39:01 | 5.79 | 2.15 | 0.039 | 1.0000 | 0.6818 | C*05:01 | 1.73 | 7.55 | 0.0034 | 0.0594 | 0.0594 |
| DRB1*15:01 | 20.72 | 8.73 | 0.0001 | 0.0013 | 0.0013 | C*15:02 | 5.63 | 11.15 | 0.0384 | 0.7301 | 0.3651 |
| DQB1*06:02 | 21.05 | 8.92 | 0.0001 | 0.001 | 0.001 | DRB1*11:01 | 16.73 | 25.45 | 0.0187 | 0.2249 | 0.1125 |
|  |  |  |  |  |  | DRB1*13:01 | 6.37 | 12.00 | 0.0347 | 0.4167 | 0.1214 |
|  |  |  |  |  |  | DRB1*14:01 | 3.59 | 8.00 | 0.0405 | 0.4857 | 0.1214 |
|  |  |  |  |  |  | DQB1*06:03 | 5.67 | 12.27 | 0.0094 | 0.0845 | 0.0422 |

\*BH-FDR: Benjamini-Hochberg False Discovery Rate

**Figure 1.**
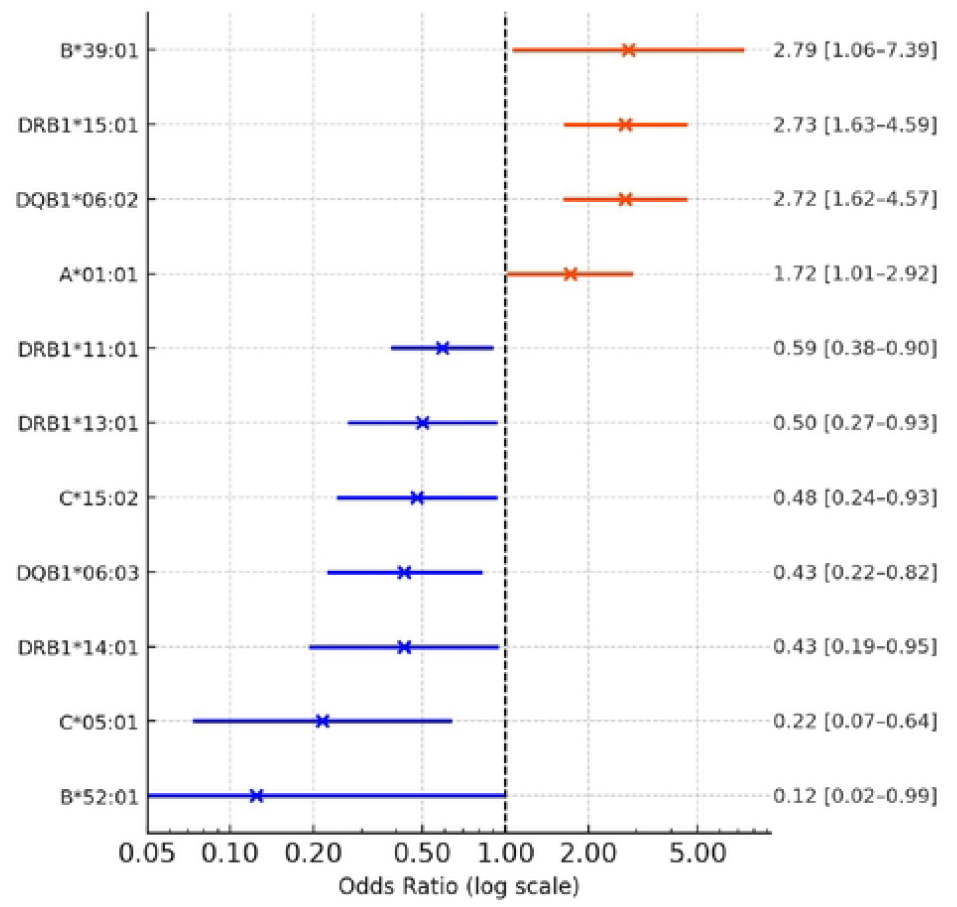
Forest plot illustrating the association between HLA alleles and multiple sclerosis in the Albanian cohort.

Notably, carrier status individuals positive for the HLA-DRB1*15:01 allele were significantly more common in MS patients (40.6%) than in controls (16.2%) (p < 0.0001; odds ratio, 3.53; 95% CI, 2.01-6.19).

Odds ratios (ORs) with 95% confidence intervals (CIs) are displayed on a logarithmic scale. The vertical dashed line indicates the null value (OR = 1.0). Alleles with confidence intervals entirely above 1.0, suggesting increased disease risk, are shown in orange-red; alleles with intervals entirely below 1.0, suggesting a protective effect, are shown in blue.

### Haplotype analysis

Five-locus haplotypes (HLA-A*∼B*∼C*∼DRB1*∼DQB1*) were inferred from the unphased genotypes of 128 patients and 148 controls using the Expectation-Maximization (EM) algorithm. The haplotypes with frequencies greater than 1% in both groups are shown in Table 3. Among patients, 17 haplotypes exceeded the 1% threshold, while only 10 haplotypes with a frequency above 1% were found in controls.

**Table 3.** Five-locus HLA haplotypes with frequencies ≥ 1% in MS patients and Controls.

| MS Patients |  |  | Controls |  |  |
| --- | --- | --- | --- | --- | --- |
| Nr . | 5-Locus Haplotype (A~B~C~DRB1~DQB1) in MS Patients | Frequency (%) | Nr . | 5-Locus Haplotype (A~B~C~DRB1~DQB1) in Controls | Frequency (%) |
| 1 | A*68:01~B*18:01~C*07:01~DRB1*15:01~DQB1*06:02 | 3.12 | 1 | A*02:01~B*18:01~C*07:01~DRB1*11:01~DQB1*03:01 | 5.41 |
| 2 | A*01:01~B*37:01~C*06:02~DRB1*16:01~DQB1*05:02 | 3.12 | 2 | A*02:01~B*51:01~C*15:02~DRB1*16:01~DQB1*05:02 | 3.38 |
| 3 | A*24:02~B*51:01~C*01:02~DRB1*04:01~DQB1*03:02 | 2.34 | 3 | A*01:01~B*08:01~C*07:01~DRB1*03:01~DQB1*02:01 | 3.04 |
| 4 | A*02:01~B*18:01~C*07:01~DRB1*11:01~DQB1*03:01 | 1.95 | 4 | A*24:02~B*35:01~C*04:01~DRB1*11:01~DQB1*03:01 | 2.99 |
| 5 | A*02:01~B*39:01~C*12:03~DRB1*12:01~DQB1*03:01 | 1.95 | 5 | A*01:01~B*37:01~C*06:02~DRB1*11:01~DQB1*03:01 | 1.69 |
| 6 | A*02:01~B*07:02~C*07:01~DRB1*15:01~DQB1*06:02 | 1.56 | 6 | A*02:01~B*51:01~C*14:02~DRB1*11:01~DQB1*03:01 | 1.35 |
| 7 | A*02:01~B*35:01~C*04:01~DRB1*15:01~DQB1*06:02 | 1.56 | 7 | A*02:01~B*51:01~C*01:02~DRB1*13:01~DQB1*06:03 | 1.35 |
| 8 | A*01:01~B*08:01~C*07:01~DRB1*03:01~DQB1*02:01 | 1.56 | 8 | A*03:01~B*07:02~C*07:01~DRB1*04:01~DQB1*03:02 | 1.01 |
| 9 | A*01:01~B*37:01~C*06:02~DRB1*11:01~DQB1*03:01 | 1.56 | 9 | A*32:01~B*40:01~C*02:02~DRB1*16:01~DQB1*05:02 | 1.01 |
| 10 | A*01:01~B*57:01~C*06:02~DRB1*04:01~DQB1*03:02 | 1.17 | 10 | A*11:01~B*35:01~C*04:01~DRB1*13:01~DQB1*06:03 | 1.01 |
| 11 | A*24:02~B*35:01~C*04:01~DRB1*10:01~DQB1*05:01 | 1.17 |  |  |  |
| 12 | A*26:01~B*47:01~C*06:02~DRB1*15:01~DQB1*06:02 | 1.17 |  |  |  |
| 13 | A*32:01~B*18:01~C*07:01~DRB1*15:01~DQB1*06:02 | 1.17 |  |  |  |
| 14 | A*01:01~B*37:01~C*06:02~DRB1*10:01~DQB1*05:01 | 1.17 |  |  |  |
| 15 | A*02:01~B*51:01~C*12:03~DRB1*11:01~DQB1*03:01 | 1.17 |  |  |  |
| 16 | A*32:01~B*35:01~C*04:01~DRB1*11:01~DQB1*03:01 | 1.17 |  |  |  |
| 17 | A*03:01~B*18:01~C*12:03~DRB1*11:01~DQB1*03:01 | 1.17 |  |  |  |

A comparative analysis identified seven haplotypes with statistically significant differences in frequency between the patient and control groups (Table 4). Of the five risk-associated haplotypes observed in the patient group, three carried the predisposing HLA-DRB1*15:01 allele, which is strongly linked to HLA-DQB1*06:02. Another haplotype carried HLA-DRB1*04:01, an allele known to be associated with autoimmune diseases.

**Table 4.** Five-locus Haplotypes with statistically significant differences between the MS patient group and the Control Group.

|  | Frequency | Frequency | *p*-value | *p*-value |
| --- | --- | --- | --- | --- |
|  | in MS | in Controls | (Fisher | (Bonferroni |
| Haplotypes | Patients (%) | (%) | nominal test) | Correction) |
| A*68:01~B*18:01~C*07:01~DRB1*15:01~DQB1*06:02 | 3.12 | 0.00 | 0.00202 | 0.0726 |
| A*24:02~B*51:01~C*01:02~DRB1*04:01~DQB1*03:02 | 2.34 | 0.00 | 0.00964 | 0.1160 |
| A*02:01~B*39:01~C*12:03~DRB1*12:01~DQB1*03:01 | 1.95 | 0.00 | 0.021 | 0.1890 |
| A*24:02~B*35:01~C*12:03~DRB1*15:01~DQB1*06:02 | 1.56 | 0.00 | 0.0457 | 0.2740 |
| A*02:01~B*07:02~C*07:01~DRB1*15:01~DQB1*06:02 | 1.56 | 0.00 | 0.0457 | 0.2350 |
| A*02:01~B*51:01~C*15:02~DRB1*16:01~DQB1*05:02 | 0.00 | 3.38 | 0.00225 | 0.0405 |
| A*02:01~B*18:01~C*07:01~DRB1*11:01~DQB1*03:01 | 1.95 | 5.41 | 0.0436 | 0.3140 |

Conversely, the haplotypes A*02:01∼B*18:01∼C*07:01∼DRB1*11:01∼DQB1*03:01 and A*02:01∼B*51:01∼C*15:02∼DRB1*16:01∼DQB1*05:02, which were most frequent in the control group, were not found among the haplotypes with a frequency greater than 1% in the patient group. Notably, the haplotype A*02:01∼B*51:01∼C*15:02∼DRB1*16:01∼DQB1*05:02 remained statistically significant after Bonferroni correction.

### Correlation of HLA-DRB1*15:01 with clinical presentation

We then examined differences in MS clinical presentations and disease severity, as defined by the various clinical forms (Relapsing-remitting, Secondary–Progressive, and Primary-Progressive) and the Expanded Disability Status Scale (EDSS), between carriers of the predisposing HLA-DRB1*15:01 allele and non-carriers. Results are presented in Table 5. No significant differences were observed in the distribution of MS clinical forms and disease severity between these groups.

**Table 5.** Correlations between clinical characteristics of patients with multiple sclerosis and the carrier status of the HLA-DRB1*15:01 allele.

| DRB1*15:01<br>allele carrier status | Age<br>(years;<br>mean,<br>SD) | Sex<br>(female;<br>nr, %) | Age at<br>disease onset<br>(years; mean,<br>SD) | Disease<br>duration<br>(years) | Relapsing -<br>remitting<br>(RR) clinical<br>form<br>(number; %) | Secondary -<br>Progressive<br>(SP) clinical<br>form<br>(number; %) | Primary -<br>Progressive<br>(PP) clinical<br>form<br>(number; %) | EDSS at<br>diagnosis | Current<br>EDSS | Relapses<br>(last 2<br>years) | MRI with<br>active lesions<br>(number; %) |
| --- | --- | --- | --- | --- | --- | --- | --- | --- | --- | --- | --- |
| DRB1*15:01<br>positive (n= 52) | 38.9<br>± 11.4 | 34<br>(65.4 %) | 29.9<br>± 9.5 | 9.0 ±<br>6.2 | 42<br>(80.8 %) | 7<br>(13.5 %) | 3<br>(5.8 %) | 2.2 ±<br>1.1 | 3.0 ±<br>1.8 | 0.7<br>± 0.8 | 32.7% |
| DRB1*15:01<br>negative (n= 76) | 37.4<br>± 12.0 | 48<br>(63.2 %) | 29.2<br>±10.9 | 8.1 ±<br>6.2 | 60<br>(78.9 %) | 7<br>(9.2 %) | 8<br>(10.5 %) | 2.0 ±<br>1.1 | 2.4 ±<br>1.9 | 0.6<br>± 0.8 | 35.5% |
| Statistical<br>significance (*p*) | 0.405 | 0.853 | 0.500 | 0.332 | 0.827 | 0.570 | 0.520 | 0.230 | 0.07 | 0.650 | 0.840 |

*EDSS*: Expanded Disability Status Scale; *MRI*: Magnetic Resonance Imaging

### Correlation between MS prevalence and HLA-DRB1 allele frequencies across Europe

To explore population-level associations, we examined the relationship between MS prevalence across 22 European populations and the corresponding allele frequencies of HLA-DRB1*15:01. As shown in Figure 2, MS prevalence was positively correlated with HLA-DRB1*15:01 frequency (Spearman’s ρ = 0.645, p = 0.0012). These findings support the role of DRB1*15:01 in the well-known north–south gradient of MS prevalence across Europe.

**Figure 2.**
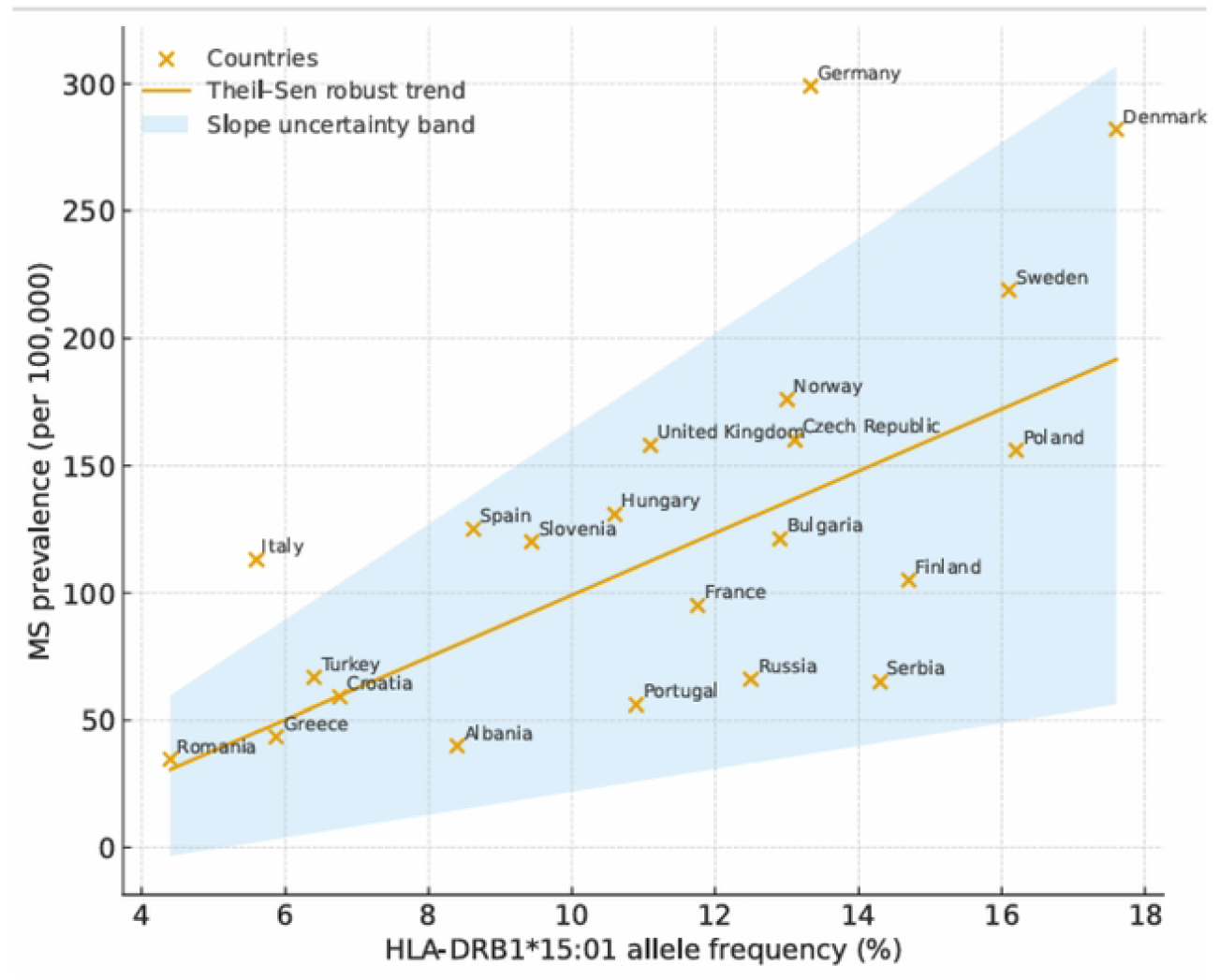
Graphical presentation of the correlation between MS prevalences (cases per 100,000 inhabitants) and *HLA-DRB1*15:01* allele frequencies (%) in 22 European countries (All data in Suppl. Table 2).

## Discussion

In this study, we explored the relationships between allele and haplotype frequencies at the HLA-A*, -B*, -C*, -DRB1*, and -DQB1* loci and multiple sclerosis (MS) in a group of Albanian patients. The DRB1*15:01 allele was the most strongly associated with MS disease compared to the general population. Other so-called predisposing alleles, such as A*01:01, B*39:01, and DQB1*06:02, were also observed at higher frequencies among patients than in the control group. However, their statistical significance was lost after Bonferroni and Benjamini–Hochberg corrections for multiple comparisons, except for DQB1*06:02. These alleles appear to have only an indirect association with MS, likely due to linkage disequilibrium within the HLA gene complex [25]. Similarly, the so-called protective alleles (B*52:01, C*05:01, C*15:02, DRB1*11:01, DRB1*13:01, DRB1*14:01, DQB1*06:03), which were more commonly found in the general population than in MS patients, also lost statistical significance after adjustments for multiple testing.

Our findings clearly show that the HLA genotype profile of Albanian MS patients differs significantly from that of the general Albanian population, a pattern also reported in other populations [16]. This difference is reflected in the more frequent presence of specific HLA haplotypes in MS patients that are rare in the general population, such as A*68:01∼B*18:01∼C*07:01-DRB1*15:01∼DQB1*06:02, which is common among MS patients but was not observed at frequencies higher than 1% in controls. Interestingly, the same haplotype has been reported as common in Ashkenazi MS patients [26].

Conversely, the haplotype A*02:01∼B*51:01∼C*15:02∼DRB1*16:01∼DQB1*05:02, which ranks as the second most common among controls, was not present among haplotypes with frequencies above 1% in MS patients. Additionally, MS patients show greater haplotype diversity than controls: 17 A*∼B*∼C*∼DRB1*∼DQB1* haplotypes were observed at frequencies above 1%, compared to only 10 in the control group.

Our study results clearly demonstrate that within the HLA system, the allele most strongly and significantly associated with MS is DRB1*15:01. The carrier status of the DRB1*15:01 allele was significantly more common in approximately 41% of MS patients compared to roughly 16% of the general population. The DQB1*06:02 allele, which also shows a strong association with MS, is linked to DRB1*15:01 through tight linkage disequilibrium [27]. Our findings in the Albanian population support numerous studies in both European and non-European populations that report the strong association between DRB1*15:01 and MS [8].

The well-known north–south gradient of MS prevalence in Europe has been widely documented [28]. We also found a direct, positive relationship between the frequency of DRB1*15:01 and MS prevalence across European populations, supporting the hypothesis that DRB1*15:01 confers a genetic predisposition to MS development, even at the population level.

The role of DRB1*15:01 in MS susceptibility has been associated with its preferential presentation of Epstein–Barr virus (EBV) epitopes in individuals who carry it. Such a presentation may trigger a pro-inflammatory autoimmune response against autoantigens that share similarities with these epitopes [29]. However, about 60% of our MS patients lack DRB1*15:01, indicating that other genetic factors outside the HLA system must also influence the disease risk [5, 30].

We also examined the potential link between DRB1*15:01 carrier status and MS clinical forms and disease severity, and found no association. Some studies have suggested that DRB1*15:01 is linked to an earlier onset and increased tissue damage, as well as a long-term worsening of symptoms [31, 32]. Still, extensive genetic studies have shown that non-HLA genes mainly influence MS severity, suggesting that DRB1*15:01 mostly affects risk rather than lifelong progression in all patients [33]. Our study has some limitations, notably the relatively small sample size in both groups, which limits the power to detect significant associations with HLA alleles other than DRB1*15:01. We also did not investigate other non-HLA genetic factors that might contribute to MS susceptibility.

In summary, our results confirm a strong association between DRB1*15:01 and MS in the Albanian population. Although it cannot serve as a definitive diagnostic marker, it may aid in diagnosis when clinical findings are unclear, especially in familiar case settings. The relatively lower frequency of this allele in the Albanian population compared to Northern Europeans, along with the higher frequency of protective alleles, may partly explain the lower MS prevalence in Albania compared to Northern Europe.

## Data Availability

All relevant data are within the manuscript and its Supporting Information files.

## Declaration of Conflicting Interests

No conflicting interests to declare

## Funding

This study was funded by the Academy of Sciences of Albania (ASA) and the National Agency for Scientific Research and Innovation (NASRI) of Albania.

**All data are available on request**.

## Notes

### Competing Interest Statement

The authors have declared no competing interest.

### Funding Statement

Yes

### Author Declarations

The study protocol received approval from the Ethics Committee of the Albanian Academy of Sciences (Project No. 373, dated July 15, 2024).

## References

1. GBD 2016 Multiple Sclerosis Collaborators. Global, regional, and national burden of multiple sclerosis 1990-2016: a systematic analysis for the Global Burden of Disease Study 2016. Lancet Neurol. 2019 Mar;18(3):269–285. doi: 10.1016/S1474-4422(18)30443-5. Epub 2019 Jan 21. PMID: 30679040; PMCID: PMC6372756.

2. Kruja J, Beghi E, Zerbi D, Dobi D, Kuqo A, Zekja I et al. High prevalence of major neurological disorders in two Albanian communities: results of a door-to-door survey. Neuroepidemiology. 2012;38(3):138–47. doi: 10.1159/000336348. Epub 2012 Mar 15. PMID: 22433124.

3. Khan Z, Mehan S, Maurya PK, Kumar A, Gupta GD, Narula AS et al. The Polygenic Nature of Multiple Sclerosis: Genetic Variants, Immunological Modulation, and Environmental Connections. Endocr Metab Immune Disord Drug Targets. 2025;25(7):527–559. doi: 10.2174/0118715303325979241206115417. PMID: 39810445; PMCID: PMC12481567.

4. Balcerac A, Louapre C. Genetics and familial distribution of multiple sclerosis: A review. Rev Neurol (Paris). 2022 Jun;178(6):512–520. doi: 10.1016/j.neurol.2021.11.009. Epub 2022 Feb 9. PMID: 35148907.

5. Ebers GC, Sadovnick AD, Dyment DA, Yee IM, Willer CJ, Risch N. Parent-of-origin effect in multiple sclerosis: observations in half-siblings. Lancet. 2004 May 29;363(9423):1773–4. doi: 10.1016/S0140-6736(04)16304-6. PMID: 15172777.

6. Koch-Henriksen N, Sørensen PS. The changing demographic pattern of multiple sclerosis epidemiology. Lancet Neurol. 2010;9(5):520–32.

7. Milo R, Kahana E. Multiple sclerosis: geoepidemiology, genetics and the environment. Autoimmun Rev. 2010;9(5):A387–94.

8. Maghbooli Z, Sahraian MA, Naser Moghadasi A. Multiple sclerosis and human leukocyte antigen genotypes: Focus on the Middle East and North Africa region. Mult Scler J Exp Transl Clin. 2020 Jan 9;6(1):2055217319881775. doi: 10.1177/2055217319881775. PMID: 31976083; PMCID: PMC6956601.

9. Bjornevik K, Cortese M, Healy BC, Kuhle J, Mina MJ, Leng Y et al. Longitudinal analysis reveals high prevalence of Epstein-Barr virus associated with multiple sclerosis. Science. 2022 Jan 21;375(6578):296–301. doi: 10.1126/science.abj8222. Epub 2022 Jan 13. PMID: 35025605.

10. Sintzel MB, Rametta M, Reder AT. Vitamin D and multiple sclerosis: a comprehensive review. Neurol Ther. 2018;7(1):59–85.

11. Wingerchuk DM. Smoking: effects on multiple sclerosis susceptibility and disease progression. Ther Adv Neurol Disord. 2012;5(1):13–22.

12. International Multiple Sclerosis Genetics Consortium (IMSGC). Multiple sclerosis genomic map implicates peripheral immune cells and microglia in susceptibility. Science. 2019;365(6460):eaav7188.

13. International Multiple Sclerosis Genetics Consortium. Multiple sclerosis genomic map implicates peripheral immune cells and microglia in susceptibility. Science. 2019 Sep 27;365(6460):eaav7188. doi: 10.1126/science.aav7188. PMID: 31604244; PMCID: PMC7241648.

14. International Multiple Sclerosis Genetics Consortium; Hafler DA, Compston A, Sawcer S, Lander ES, Daly MJ, De Jager PL et al. Risk alleles for multiple sclerosis identified by a genomewide study. N Engl J Med. 2007 Aug 30;357(9):851–62. doi: 10.1056/NEJMoa073493. Epub 2007 Jul 29. PMID: 17660530.

15. Gregory SG, Schmidt S, Seth P, Oksenberg JR, Hart J, Prokop A et al. Multiple Sclerosis Genetics Group. Interleukin 7 receptor alpha chain (IL7R) shows allelic and functional association with multiple sclerosis. Nat Genet. 2007 Sep;39(9):1083–91. doi: 10.1038/ng2103. Epub 2007 Jul 29. PMID: 17660817.

16. Hauser SL, Fleischnick E, Weiner HL, Marcus D, Awdeh Z, Yunis EJ et al. Extended major histocompatibility complex haplotypes in patients with multiple sclerosis. Neurology. 1989 Feb;39(2 Pt 1):275–7. doi: 10.1212/wnl.39.2.275. PMID: 2783768.

17. Ferrè L, Filippi M, Esposito F. Involvement of Genetic Factors in Multiple Sclerosis. Front Cell Neurosci. 2020 Dec 1;14:612953. doi: 10.3389/fncel.2020.612953. PMID: 33335478; PMCID: PMC7735985.

18. Thompson AJ, Banwell BL, Barkhof F, Carroll WM, Coetzee T, Comi G et al. Diagnosis of multiple sclerosis: 2017 revisions of the McDonald criteria. Lancet Neurol. 2018 Feb;17(2):162–173. doi: 10.1016/S1474-4422(17)30470-2. Epub 2017 Dec 21. PMID: 29275977.

19. Ikhtiar AM, Jazairi B, Khansa I, Othman A. HLA class I alleles frequencies in the Syrian population. BMC Res Notes. 2018 May 21;11(1):324. doi: 10.1186/s13104-018-3427-1. PMID: 29784010; PMCID: PMC5963146.

20. Excoffier L, Laval G, Schneider S. Arlequin ver. 3.5: an integrated software package for population genetics data analysis. Mol Ecol Resour. 2010;10(3):564–567. doi:10.1111/j.1755-0998.2010.02847.x

21. Bland JM, Altman DG. Multiple significance tests: the Bonferroni method. BMJ. 1995;310(6973):170. doi:10.1136/bmj.310.6973.170.28

22. Benjamini Y, Hochberg Y. Controlling the false discovery rate: a practical and powerful approach to multiple testing. J R Stat Soc Series B. 1995;57(1):289–300.

23. González-Galarza FF, Takeshita LY, Santos EJ, Kempson F, Maia MH, da Silva AL et al. Allele frequency net 2015 update: new features for HLA epitopes, KIR and disease and HLA adverse drug reaction associations. Nucleic Acids Res. 2015 Jan;43(Database issue):D784-8. doi: 10.1093/nar/gku1166. Epub 2014 Nov 20. PMID: 25414323; PMCID: PMC4383964.

24. Multiple Sclerosis International Federation (MSIF). New prevalence and incidence data now available in the Atlas of MS [Internet]. Aug 21 2023 [cited Sep 10 2025]. Available from: atlasofms.org.

25. Chao MJ, Barnardo MC, Lui GZ, Lincoln MR, Ramagopalan SV, Herrera BM et al. Transmission of class I/II multi-locus MHC haplotypes and multiple sclerosis susceptibility: accounting for linkage disequilibrium. Hum Mol Genet. 2007 Aug 15;16(16):1951–8. doi: 10.1093/hmg/ddm142. Epub 2007 Jun 20. PMID: 17584771.

26. Khankhanian P, Matsushita T, Madireddy L, Lizée A, Din L, Moré JM et al. Genetic contribution to multiple sclerosis risk among Ashkenazi Jews. BMC Med Genet. 2015 Jul 28;16:55. doi: 10.1186/s12881-015-0201-2. PMID: 26212423; PMCID: PMC4557862.

27. De Silvestri A, Capittini C, Mallucci G, Bergamaschi R, Rebuffi C, Pasi A et al. The Involvement of HLA Class II Alleles in Multiple Sclerosis: A Systematic Review with Meta-analysis. Dis Markers. 2019 Nov 6;2019:1409069. doi: 10.1155/2019/1409069. PMID: 31781296; PMCID: PMC6875418.

28. Compston A. The story of multiple sclerosis. In: A Compston, G Ebers, H Lassman, et al. (eds) McAlpine’s multiple sclerosis. London: Churchill Livingstone, 1998, pp.3–44.

29. Tschochner M, Leary S, Cooper D, Strautins K, Chopra A, Clark H et al. Identifying Patient-Specific Epstein-Barr Nuclear Antigen-1 Genetic Variation and Potential Autoreactive Targets Relevant to Multiple Sclerosis Pathogenesis. PLoS One. 2016 Feb 5;11(2):e0147567. doi: 10.1371/journal.pone.0147567. PMID: 26849221; PMCID: PMC4744032.

30. Shirai R, Yamauchi J. New Insights into Risk Genes and Their Candidates in Multiple Sclerosis. Neurol Int. 2022 Dec 29;15(1):24–39. doi: 10.3390/neurolint15010003. PMID: 36648967; PMCID: PMC9844300.

31. Brownlee WJ, Tur C, Manole A, Eshaghi A, Prados F, Miszkiel KA et al. HLA-DRB1*1501 influences long-term disability progression and tissue damage on MRI in relapse-onset multiple sclerosis. Mult Scler. 2023 Mar;29(3):333–342. doi: 10.1177/13524585221130941. Epub 2022 Nov 18. PMID: 36398585.

32. Tur C, Ramagopalan S, Altmann DR, Bodini B, Cerchignani M, Khaleeli Z. et al. HLA-DRB1*15 influences the development of brain tissue damage in early PPMS. Neurology. 2014;83(19):1712–1718. doi:10.1212/WNL.0000000000000959.

33. International Multiple Sclerosis Genetics Consortium; MultipleMS Consortium. Locus for severity implicates CNS resilience in progression of multiple sclerosis. Nature. 2023 Jul;619(7969):323–331. doi: 10.1038/s41586-023-06250-x. Epub 2023 Jun 28. PMID: 37380766; PMCID: PMC10602210.

